# Detection of COVID-19 Outbreaks in Long-Term Care Homes Using Built Environment Testing for SARS-CoV-2: A Multicentre Prospective Study

**DOI:** 10.1101/2022.06.28.22276560

**Authors:** Michael Fralick, Caroline Nott, Jason Moggridge, Lucas Castellani, Ashley Raudanskis, David S. Guttman, Aaron Hinz, Nisha Thampi, Alex Wong, Doug Manuel, Allison McGeer, Evgueni Doukhanine, Hebah Mejbel, Veronica Zanichelli, Madison Burella, Sylva Donaldson, Pauline W. Wang, Rees Kassen, Derek MacFadden

## Abstract

**Background:** Environmental surveillance of SARS-CoV-2 via wastewater has become an invaluable tool for population-level surveillance of COVID-19. Built environment sampling may provide a more spatially refined approach for surveillance of COVID-19 in congregate living settings and other high risk settings (e.g., schools, daycares).

**Methods:** We conducted a prospective study in 10 long-term care homes (LTCHs) across three cities in Ontario, Canada between September 2021 and May 2022. Floor surfaces were sampled weekly at multiple locations (range 10 to 24 swabs per building) within each building and analyzed for the presence of SARS-CoV-2 using RT-qPCR. The exposure variable was detection of SARS-CoV-2 on floors. The primary outcome was the presence of a COVID-19 outbreak in the week that floor sampling was performed.

**Results:** Over the 9-month study period, we collected 3848 swabs at 10 long-term care homes. During the study period, 19 COVID-19 outbreaks occurred with 103 cumulative weeks under outbreak. During outbreak periods, the proportion of floor swabs positive for SARS-CoV-2 was 50% (95% CI: 47-53) with a median quantification cycle of 37.3 (IQR 35.2-38.7). During non-outbreak periods the proportion of floor swabs positive was 18% (95% CI:17-20) with a median quantification cycle of 38.0 (IQR 36.4-39.1). Using the proportion of positive floor swabs for SARS-CoV-2 to predict COVID-19 outbreak status in a given week, the area under the receiver operating curve (AUROC) was 0.85 (95% CI: 0.78-0.92). Using thresholds of ≥10%, ≥30%, and ≥50% of floor swabs positive for SARS-CoV-2 yielded positive predictive values for outbreak of 0.57 (0.49-0.66), 0.73 (0.63-0.81), and 0.73 (0.6-0.83) respectively and negative predictive values of 0.94 (0.87-0.97), 0.85 (0.78-0.9), and 0.75 (0.68-0.81) respectively. Among 8 LTCHs with an outbreak and swabs performed in the antecedent week, 5 had positive floor swabs exceeding 10% at least five days prior to outbreak identification. For 3 of these 5 LTCHs, positivity of floor swabs exceeded 10% more than 10 days before the outbreak being identified.

**Conclusions:** Detection of SARS-CoV-2 on floors is strongly associated with COVID-19 outbreaks in LTCHs. These data suggest a potential role for floor sampling in improving early outbreak identification.

## INTRODUCTION

The COVID-19 pandemic has disproportionately affected older frail adults, particularly residents of long-term care homes (LTCHs). At the beginning of the COVID-19 pandemic, mortality from COVID-19 during outbreaks in LTCHs exceeded 30% in many regions.^1,2^ The high mortality reflected the rapidity with which SARS-CoV-2 could spread in congregate settings, the vulnerability of the residents, and insufficient infection control measures. Even after the development of vaccines and an improved understanding of transmission dynamics, COVID-19 outbreaks continue to occur in LTCHs.^3^ One key element in outbreak prevention is consistent and proactive disease surveillance, particularly in high-risk congregate living settings such as LTCHs. The primary method of surveillance during the pandemic has included “active” approaches such as clinical testing of individuals, though this is costly and difficult to sustain. A passive surveillance method is environmental detection of SARS-CoV-2, and this might be one approach for non-invasive surveillance that could be applied in congregate settings.

The primary environmental surveillance approach to-date for COVID-19 has been wastewater surveillance.^4-7^ Wastewater surveillance is a helpful public-health tool because it is a passive, non-invasive, and can detect early transmission of SARS-CoV-2 in the community. However, it lacks standardization, reproducibility, and the spatial resolution as it is typically reported on a regional-level. And while wastewater sampling can be performed on individual buildings (e.g., University Residence^8,9^) and has been shown to be actionable to mitigate transmission, this is not generalizable to LTCHs for two main reasons. First, many residents of LTCHs don’t use the toilet due to incontinence. Second, the costs, logistics, and necessary work to establish wastewater surveillance in LTCHs can be prohibitive. Testing individuals, on the other hand, faces limitations around cost, performance, accessibility and acceptance. We need to bridge the gap between population-level surveillance methods like wastewater testing and individual-level surveillance methods. Recent studies have demonstrated that SARS-CoV-2 can be recovered from the built environment, in particular floors.^10-12^

SARS-CoV-2 can remain viable on stainless steel and plastics for approximately 6 hours and its RNA can be detected for days thereafter.^13^ Despite detection on surfaces, fomite transmission of SARS-CoV-2 through direct contact is uncommon.^14^ Transmission via respiratory particles often occurs when a person is asymptomatic, pre-symptomatic, or minimally symptomatic,^15^ making early and reliable identification of transmission challenging, particularly in the LTCH setting.^16^ Where individual-level screening is costly, resource intensive, and invasive, environmental surface sampling has the potential to provide timely detection and localize cases to a physical space. Floors can act as a “sink” for respiratory viral particles produced by infected individuals. We have previously shown a strong correlation between the presence of SARS-CoV-2 RNA from floors and patient-level COVID-19 burden in acute care hospital settings.^10^ We propose that floor swabbing could be used to detect outbreaks of COVID-19 in LTCHs, representing a potential new tool for early identification of outbreaks. Our objective was to assess whether the presence of SARS-CoV-2 RNA on the floors of the built environment was associated with COVID-19 outbreaks.

## Methods

### Study Design

We conducted a multicentre prospective study in a convenience sample of 10 LTCHs in Ontario, Canada, between September 2021 and May 2022. In Ontario, LTCHs are administered by the Ministry of Long-term Care and are a place of residence for older adults. The residents receive both personal care and nursing care in addition to subsidized accommodation under a publicly funded government LTCH program.^17^ LTCHs were located in large [Toronto (n=3) and Ottawa (n=5)] and small [Sault Ste Marie (n=2)] urban settings. The LTCH staff were blinded to swab results for at least 8 weeks after the swabs were performed and processed. COVID-19 outbreak and weekly case count data were reported to us by the LTCH managers and cross-checked with public health records.^18^ Up until April 2022, all of the study sites required staff and visitors to perform a rapid antigen test upon entry to the home, wear surgical masks while in the home, and perform regular hand hygiene.^19^ Some sites also required staff members to wear a face shield as well. In addition, there was weekly testing for staff. Staff members with a positive rapid antigen or PCR test were immediately sent home from work. Our study was reviewed by the research ethics board at the University of Ottawa and received a waiver because the sample collected are not human samples (see Article 2.1 of the TCPS 2), and any demographic data is either a) publicly available through a mechanism set out by legislation or regulation and that is protected by law; or b) in the public domain and the individuals to whom the information refers have no reasonable expectation of privacy (see Article 2.2.).

### Swabbing locations

At each LTCH, we had a site visit with the LTCH manager and their infection control team to identify swab locations. The swab locations included common areas shared by both residents and staff (e.g., dining areas, recreation rooms, hallways) and staff-only areas (e.g., staff locker rooms, staff lunch rooms, kitchen, laundry areas). Swab samples were taken from the same locations weekly. For privacy reasons we did not swab within any of the residents’ rooms. Between 10-20 swabs were performed each week at each LTCH depending on the size of the building.

### Swabbing procedure and processing

Floor swabs were collected by either LTCH staff (Ottawa) or study personnel (Toronto and Sault Ste Marie). Sample collection followed previously validated protocols^10^, and involved initial swab wetting in nucleic acid stabilization solution followed by approximately 30 seconds of swabbing across a 2”x 2” area. Floors were sampled using the P-208 Environmental Surface Collection Prototype kit from DNA Genotek. The kit consists of a flocked swab and 2 mL of semi-lytic nucleic acid stabilization solution for post-collection swab immersion. The swabs were then sent to our lab and SARS-CoV-2 was detected by quantitative reverse-transcriptase polymerase chain reaction (RT-qPCR) of RNA extracted from the stabilization solution using the MagMAX Viral/Pathogen II (MVP II) Nucleic Acid Isolation Kit (Thermo Fisher Scientific, Waltham, MA). Our previous work provides in depth information regarding the validation of swabbing of the built environment for SARS-CoV-2 detection.^10^ The RT-qPCR results provided a quantification cycle (Cq) of detection for each positive swab. For our study we considered a positive result to be a Cq value less than 45, consistent with cut-offs used for wastewater surveillance and other qPCR applications.^8^

### Data Sources

In addition to the RT-qPCR SARS-CoV-2 detection, we also collected carbon dioxide (CO_2_) measurements for a subset of homes (Toronto and Sault Ste. Marie, n=5) and other covariates related to the built environment. CO_2_ measurements were collected using the Indoor Air Quality 9,999 parts per million (ppm) Digital Carbon Dioxide Temperature Humidity NDIR Sensor. CO_2_ levels are an indicator of ventilation and have been proposed as a means to estimate risk of COVID-19 infection in an indoor space.^20^ The CO_2_ measurements were taken at the same locations were the swabs were performed. Other covariates included information related to the building (year of construction, number of floors, number of rooms, number of single and multi-occupancy rooms, number of residents, number of staff), staff (percentage fully vaccinated at the start of the study), residents (percentage fully vaccinated), ventilation system, information on past outbreaks, and infection control measures (cleaning protocols, the type of personal protective equipment worn by staff and residents, and whether or not PCR and/or rapid COVID-19 tests are required for workers and visitors).^19^ Data on the number of COVID-19 cases at each facility during our study period were collected in aggregate and we did not use individual identifying data. We did, however, obtain the number of infected individuals who were residents versus staff members. We used provincially reported outbreak case data^18^ to ensure that all LTCH outbreaks were captured in our dataset, however for specific dates of outbreaks we used LTCH reported values as they were felt to be most reflective of actual practices within the homes.

### Study Outcomes

The primary outcome was the presence of a COVID-19 outbreak in a LTCH during a given week. Outbreak status was as reported by the LTCH, which is typically defined according to provincial guidance in Ontario set prior to the start of this study, namely “..an outbreak is defined as two or more lab-confirmed COVID-19 cases in residents, staff or other visitors in a home, with an epidemiological link, within a 14-day period, where at least one case could have reasonably acquired their infection in the LTC home.”^18^ We used case data collected from the LTCHs, and validated these with provincially reported data as noted above.^18^ As a secondary outcome we evaluated individual staff-level cases where an outbreak did not occur.

### Statistical Analysis

Descriptive statistics are reported, including counts (outbreaks), proportions (positive swabs), and continuous values (quantitation cycle and CO_2_ measurements). Statistical comparison of PCR quantification cycle (Cq) values was performed using a Mann-Whitney U test, given their non-normal distribution (by Shapiro-Wilk tests). Confidence intervals were calculated for proportions using Wilson’s method. We determined the test characteristics of the proportion of PCR-positive swabs for predicting an outbreak in a given facility for a given week. The sensitivity, specificity, negative predictive value (NPV), and positive predictive value (PPV), of the proportion of PCR-positive swabs were estimated for various decision thresholds. Similarly, we created a logistic regression model for outbreak detection using the proportion of PCR-positive swabs as predictor; where the AUROC was estimated by bootstrapping and evaluating predictions for each out-of-bag sample (*B*=2000, percentile method). We stratified outcomes by location type, including shared and worker areas. We also quantified how the percentage of positive swabs changed in the weeks leading up to, and following, an outbreak being identified by the LTCH. To assess the relationship between CO_2_ and the detection of SARS-CoV-2 we compared the CO_2_ concentrations observed at collection for positive and negative swabs. All statistical analyses were performed using R (version 4.2).

## RESULTS

Over a 9-month time period we serially swabbed 10 LTCHs. Summary statistics for all covariates are given in Table 1. All sites had vaccination rates exceeding 95% for both staff and residents. Among the LTCHs, most buildings (70%) were built prior to 1980, 2 had primarily single occupancy rooms, and there were an average of 155 residents per LTCH and 213 staff per LTCH. A total of 3,848 swabs were collected (mean 385 per home), 2,024 CO_2_ measurements were recorded (50% of LTCHs), and 19 COVID-19 outbreaks were declared.

**Table 1.** Baseline characteristics of the included long-term care homes.

| N homes | 10 |
| --- | --- |
| **Buildings** |  |
| Year built | 1974 (1966, 2004) |
| Number of floors | 3 (1, 7) |
| Total rooms | 125 (58, 533) |
| Total residence rooms | 112 (35, 374) |
| % single occupancy | 29.6 (5.3, 100) |
| **Resident level** |  |
| Number of residents | 149.5 (63, 300) |
| Percentage vaccinated | 98.15 (92.8, 100) |
| **Staff level** |  |
| Number of staff | 200.5 (93, 383) |
| Percentage vaccinated | 100 (95, 100) |
| Expressed as *median (min, max)* |  |

The median duration of an outbreak was 35 days and 18 (95%) involved both residents and workers, with the median number of cases being 34 (range: 2-150). For swabs performed during outbreak periods, the prevalence of positive swabs was 50% (47-53), with a median Cq of 37.3 (IQR 35.2-38.7) (table 2). For swabs performed during non-outbreak periods, the prevalence of positive swabs was 18% (17-20), with a median Cq of 38.0 (IQR 36.4-39.1). The median Cq of swabs during non-outbreak periods was significantly lower (p<0.01), indicating a higher quantity of viral RNA sampled, during outbreak periods compared to non-outbreak periods. No outbreaks only involved staff and not residents.

**Table 2.** Detection of SARS-CoV2 and CO_2_ readings stratified by outbreak status

|  | Overall | Outbreak | Staff cases | No cases |
| --- | --- | --- | --- | --- |
| **Percentage of PCR-positive swabs** |  |  |  |  |
| All locations | 28.9 (27.5-30.4) | 50.1 (47.4-52.8) | 12.8 (9.4-17.1) | 18.9 (17.3-20.6) |
| Shared areas* | 28.6% (26.8-30.4) | 48.9% (45.5-52.2) | 8.7% (5.7-13) | 19.9% (17.8-22) |
| Worker-only areas | 29.5% (27.1-32) | 52.3% (47.7-56.8) | 28.8% (18.8-41.4) | 17.2% (14.7-19.9) |
| **Cycle threshold of positive swabs** |  |  |  |  |
| Cycle threshold | 37.7 (35.6-38.9) | 37.3 (35.2-38.7) | 38 (36.4-38.7) | 38 (36.3-39.1) |
| **Covariates** |  |  |  |  |
| Carbon-dioxide (ppm) | 522 (458-597) | 523 (473-586) | 424 (376-499) | 536 (471-612) |

PCR-positive swabs expressed as percentage with 95% CI by Wilson method
Cycle-threshold and CO<sub>2</sub> expressed as median (IQR)
\* refers to areas in the long-term care home where the space is shared by both residents and workers

For swabs performed during periods with identified staff cases (within 72 hours of case detection) but no resident-level cases or outbreaks, the prevalence of positive swabs was 12.8% (9.4-17.1), and with a median Cq of 38 (IQR 36.4-38.7). Four LTCHs had staff cases that did not lead to an outbreak and when there were staff cases the swab positivity was 28.8% (IQR 18.8-41.4) in worker-only areas (e.g., locker rooms, staff dining areas) compared to 8.7% (5.7%-13%) for swabs performed in areas that included both staff and residents.

The percentage of positive swabs increased in the days and weeks prior to an outbreak being identified by the LTCH (Table 3). The percentage of positive swabs also decreased in the days and weeks after an outbreak was identified by the LTCH (Table 3). Among eight LTCHs with an outbreak and swabs performed in the weeks prior to the outbreak starting, five of the homes had floor swab positivity exceeding 10% five or more days prior to the outbreak being identified (Figure 1). For three of these five LTCHs, positivity of floor swabs exceeded 10% more than 10 days before the outbreak was identified. We generated receiver operating characteristic (ROC) curves for the primary outcome (presence of outbreak in a given week) using the proportion of swabs positive for SARS-CoV-2 and this yielded an area under the ROC of 0.85 (95% CI: 0.78-0.92). The PPV, NPV, specificity and sensitivity are provided in Table 4 across five thresholds of swab positivity (i.e., 10%, 20%, 30%, 40%, and 50%) and displayed graphically across all thresholds (Figure 2). The percentage of positive swabs was not associated with CO_2_ levels in PPM (Appendix).

**Table 3.** Percentage of swabs positive for SARS-CoV2 in the days leading up to the identification of an outbreak and after the end of an outbreak.

|  | Days before outbreak |  |  | Days after outbreak |  |
| --- | --- | --- | --- | --- | --- |
|  | 15:21 | 8:14 | 1:7 | +1:7 | +8:14 |
| n Swabs | 265 | 238 | 279 | 161 | 183 |
| % Positive | 23% (19-29) | 34% (28-40) | 42% (37-48) | 48% (40-56) | 36% (29-43) |
| Cycle threshold | 37.6 (35.8-38.8) | 38.7 (36.6-39.5) | 37.5 (34.9-38.7) | 37.7 (36.4-39.3) | 38.4 (36.5-39.2) |

% positive swabs expressed as *percentage* (95% CI).
CIs computed by Wilson's method for binomial proportions.
PCR cycle threshold values expressed as *median* (IQR).

**Table 4.** Test characteristics of floor swabs to identify a current outbreak.

| Threshold | Sensitivity | Specificity | NPV | PPV |
| --- | --- | --- | --- | --- |
| 10% | 0.93 (0.85-0.97) | 0.6 (0.52-0.68) | 0.94 (0.87-0.97) | 0.57 (0.49-0.66) |
| 20% | 0.84 (0.75-0.91) | 0.72 (0.64-0.79) | 0.89 (0.82-0.93) | 0.64 (0.54-0.72) |
| 30% | 0.75 (0.64-0.83) | 0.84 (0.77-0.89) | 0.85 (0.78-0.9) | 0.73 (0.63-0.81) |
| 40% | 0.6 (0.49-0.7) | 0.86 (0.8-0.91) | 0.79 (0.72-0.85) | 0.71 (0.6-0.81) |
| 50% | 0.49 (0.39-0.6) | 0.9 (0.84-0.94) | 0.75 (0.68-0.81) | 0.73 (0.6-0.83) |
| Test characteristics expressed as estimate (95% CI by Wilson) |  |  |  |  |

**Figure 1.**
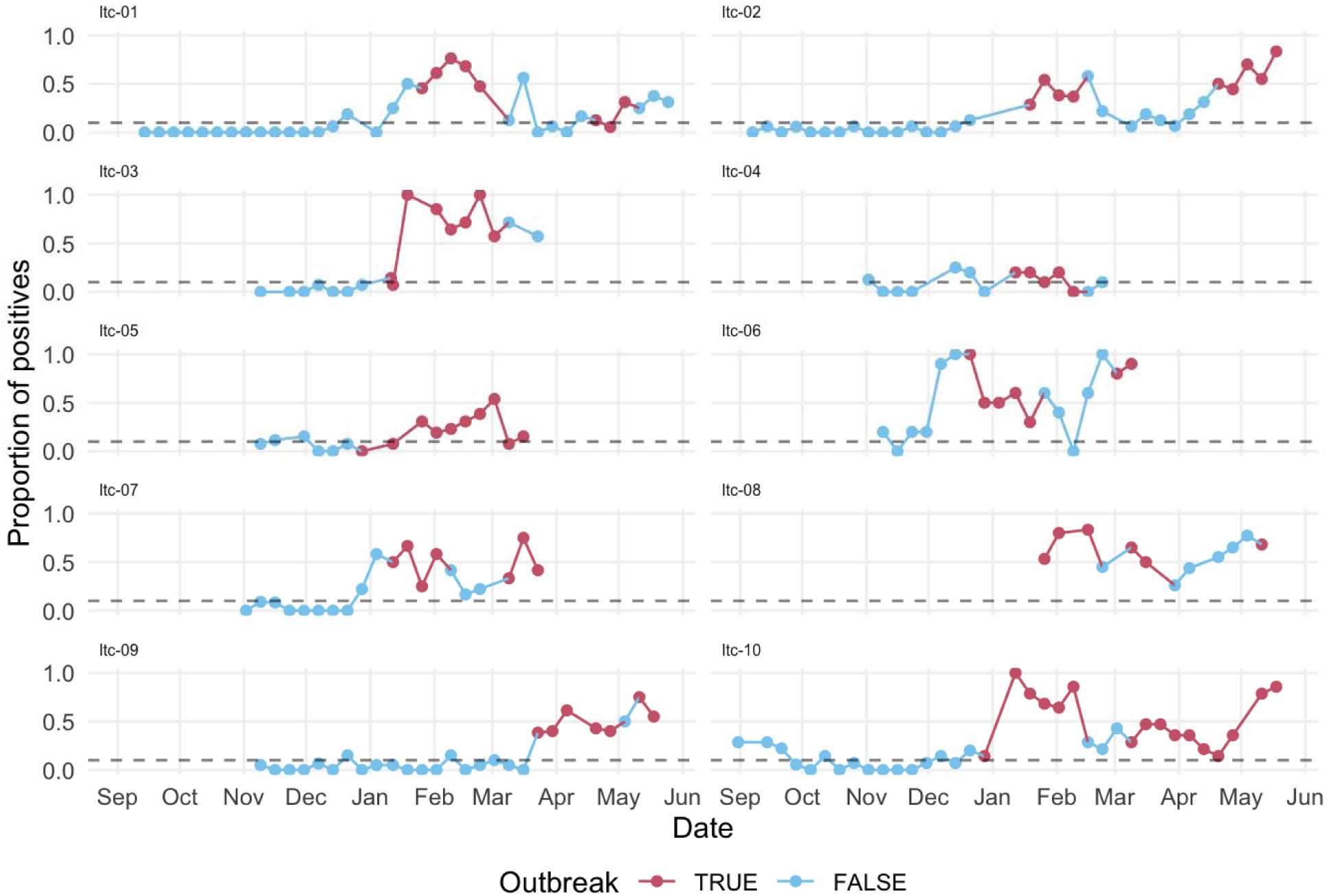
A graphical representation of swab positivity for SARS-CoV2 over time Legend - Red lines depict time periods when the long-term care home identified an outbreak was present and blue lines depict periods when there was no outbreak identified by the long-term care home. Dashed line represents a threshold of 10% of swabs positive for SARS-CoV-2.

**Figure 2.**
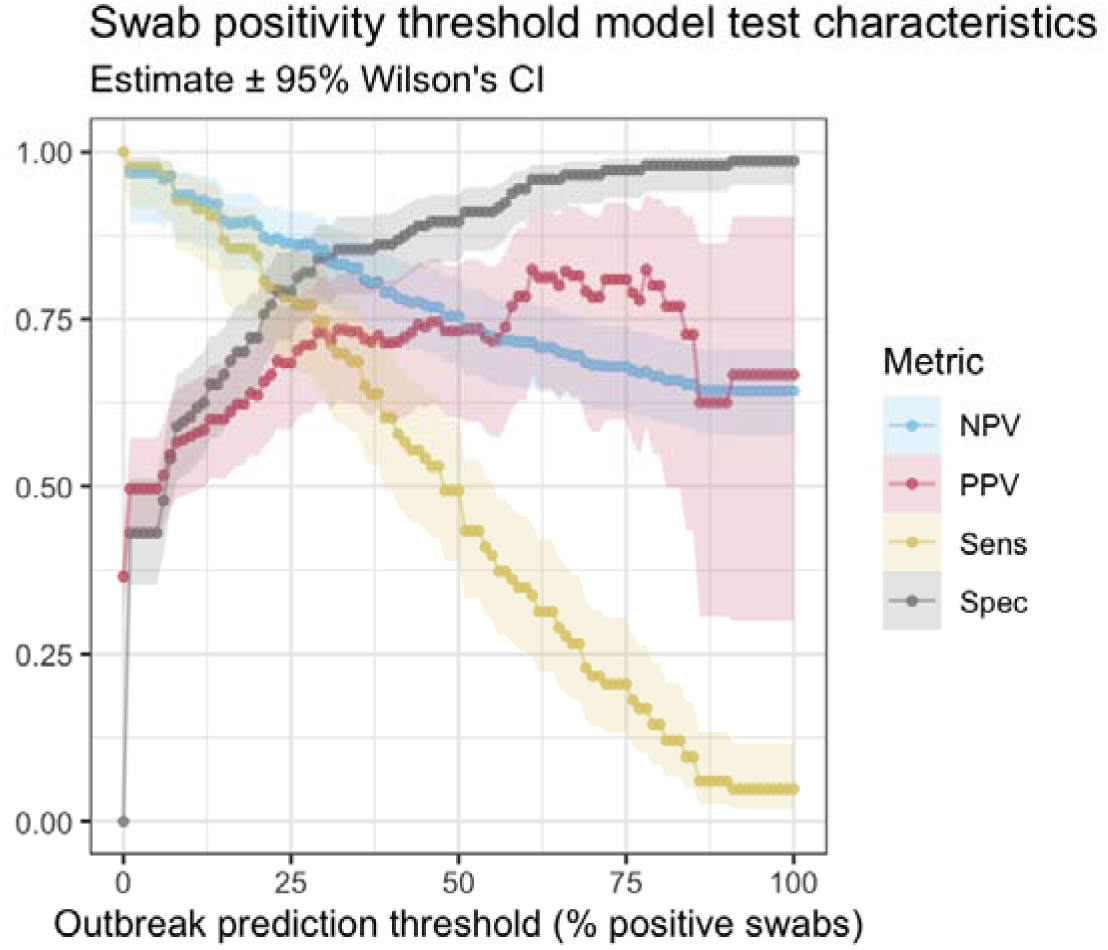
Test characteristics across swab positivity ranging from 0% to 100% Legend. NPV = negative predictive value, PPV = positive predictive value, Sens = sensitivity, Spec = specificity. Estimates are shown as connected points with shading for the 95% CI range.

For swabs performed during periods with no identified staff or resident-level cases and no outbreak, the prevalence of positive swabs was 18.9% (17.3-20.6), with a median Cq of 38 (IQR 36.3-39.1).

## DISCUSSION

In this multicenter prospective study, we found that detection of SARS-CoV-2 from the built environment was strongly associated with COVID-19 outbreaks. Furthermore, the rising proportion of positive floor swabs preceded the formal recognition of an outbreak by at least 5 days in a significant fraction of outbreaks. In addition, the prevalence of positive floor swabs for SARS-CoV-2 also declined after the cessation of outbreaks. These results suggest that built environment SARS-CoV-2 testing could be a valuable tool for identifying and monitoring of SARS-CoV-2 outbreaks in LTCHs.

Since the emergence and dominance of the Omicron variant, many jurisdictions globally have shifted from individual level testing to wastewater monitoring as a means for population-level COVID-19 surveillance.^4-7^ Wastewater surveillance allows for the quantitation of SARS-CoV-2 within waste effluent, a metric that is correlated with the burden of disease in the population of the waste catchment.^21-23^ While the results can provide surveillance on a population-level, one of the major criticisms is that they generally cannot provide results that are directly actionable. Built environment sampling of floors has high accuracy, likely because SARS-CoV-2 spreads via respiratory particles that eventually settle to the floor surface where they can be detected.^16^ Because people can spread the virus during pauci- or asymptomatic states, sampling the built environment could provide a localized advanced warning signal. Our study supports this assertion and suggests that surface sampling can provide data to identify an impending (or clinically undetected) outbreak and possibly also aid in monitoring whether an outbreak is worsening, improving, or resolved. As a result, swabbing the built environment is likely complementary to wastewater surveillance and the importance of these approaches will rise as infection control measures continue to be relaxed in congregate settings.

There are many ways in which our results could direct infection prevention and control activities in LTCHs. For example, if high levels of swab positivity are only detected in worker-facing areas (e.g., locker rooms) then it can inform the need to specifically test workers. If high levels of swab positivity are in resident-facing areas then it can potentially inform infection control measures such as universal masking among residents and visitors, cohorting and testing residents, and influence visitor policies. In doing so, there is the potential for early identification of an outbreak before it occurs which can drive actions (e.g. focused human testing, additional PPE and isolation requirements) that are known to either prevent or mitigate further transmission. Considering the high rate of morbidity and mortality of outbreaks in LTCHs, this has the potential to save lives. In addition, there is the potential that this approach could also prevent unnecessary enactment of isolation measures in certain circumstances (e.g. staff cases with low floor signal indicating lack of ongoing transmission amongst residents).

Another important reason for early identification of outbreaks among high risk populations is the initiation of early treatment. There are multiple treatments available for high-risk populations including both oral (e.g., nirmatrelvir/ritonavir) and intravenous (e.g., remdesivir) medications.^24,25^ Both nirmatrelvir/ritonavir and remdesivir reduce a person’s relative risk of hospitalization or death by approximately 90%. However, these medications are only effective if given within days of symptom onset which provides added importance of early identification of cases within an LTCH.^24,25^ Future studies will be needed to identify if swabbing floors will indeed lead to earlier diagnosis of COVID-19 among residents and staff, with subsequent access to treatments where appropriate.

There are important limitations of our study. First the public health definition of “outbreak” has inherent limitations because the ideal goal of swabbing is to know whether there is the potential (or evidence for) ongoing SARS-CoV-2 transmission in a given setting, and public health definition of two or more epidemiologically linked (and clinically detected) cases is neither perfectly sensitive nor specific for this. We attempted to better understand the potential utility of floor swabbing by analyzing how well the swab results correlate with reported cases that have not reached outbreak status, and our results suggest that swabbing the floor is sensitive enough to detect even a single infected worker. Second, we included long-term care homes that represent a convenience sample rather than a random sample, and thus our results might not generalize to all long-term care homes, though we purposefully included long-term care homes across multiple cities and settings (urban/rural) to improve generalizability. Third, our study showed a clear association between the detection of SARS-CoV-2 on floors and an impending outbreak but it is unknown how providing this information in real-time could change the course of outbreaks. Fourth, at most sites we swabbed once per week and thus it is unknown if swabbing more often might improve the timeliness of our outbreak identification.

One important future direction of our work is the assessment for other pathogens in not only LTCHs, but also hospitals and other congregate settings. For LTCHs, both respiratory outbreaks and diarrhea illness are common and thus identifying the presence of such pathogens (e.g., influenza, respiratory syncytial virus, norovirus) may prove useful. Wastewater surveillance has recently moved to expand to include some of these pathogens, and highlights how our approach can be complementary, including the potential to detect pathogens not shed in stool. Finally, early detection of emerging pathogens (e.g., monkey pox) is a natural potential extension of our work and is an area of active investigation.

Our multicentre prospective study of long-term care homes demonstrates that SARS-CoV-2 detection in the built environment is highly correlated with human-level cases and COVID-19 outbreaks, and environmental detection often precedes the identification of an outbreak. Swabbing the built environment, similar to wastewater surveillance, may provide another tool for infection prevention and control for combating COVID-19 in congregate settings. Future studies evaluating the implementation and generalizability of this approach are needed.

## Data Availability

All data produced in the present work are contained in the manuscript.

## Acknowledgements

We thank Natasha Milijasevic, Cory Nezan, Dr. Matthew Morgan, and the Extendicare long-term care homes that participated in our study. We also thank Marion Godoy and the Responsive Health Management Incorporated long-term care homes that participated in our study. Finally, we thank Dr. Ed Hirvi and FJ Dave Long Term Care Home and, Dr. Lisa Perry and the team at the Ontario Finnish Rest Home.

## APPENDIX

**Appendix Table 1.**
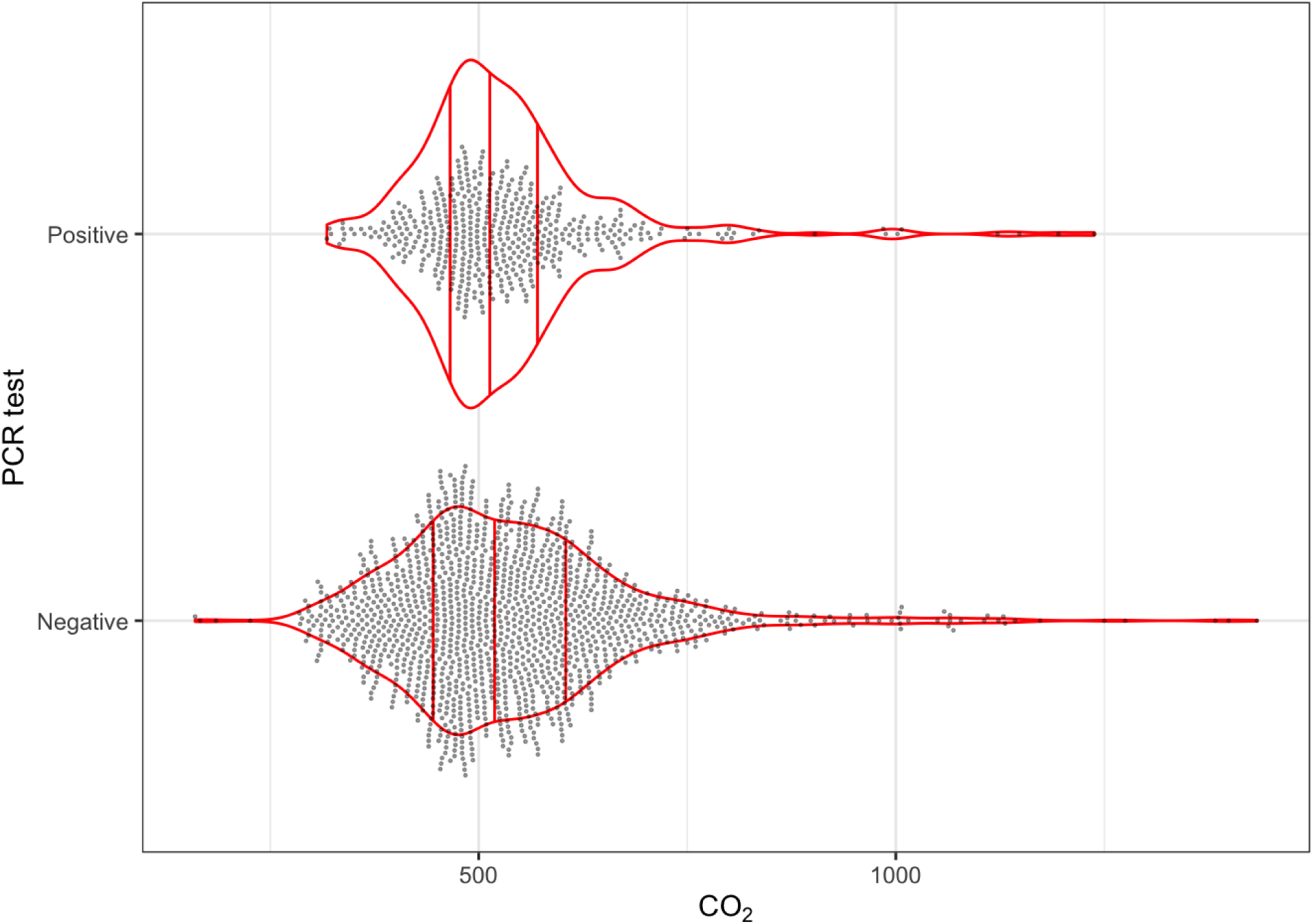
A plot of the association between CO2 and the PCR test result of the floor swab

